# Frequent gastrointestinal cancer complications in Japanese patients with coronary syndromes undergoing percutaneous coronary intervention

**DOI:** 10.1101/2024.12.17.24319200

**Authors:** Yasuyuki Chiba, Shogo Imagawa, Yuki Takahashi, Kimitoshi Kubo, Kenta Otsuka, Kyo Shimazu, Teisuke Anzai, Kazuya Yonezawa, Mototsugu Kato, Toshihisa Anzai

**Author notes:** Address for correspondence: Dr. Yasuyuki Chiba, Division of Cardiology, National Hospital Organization Hakodate Medical Center, 18-16, Kawahara-cho, Hakodate, Hokkaido 041-8512, Japan.

## Abstract

**Background:** Gastrointestinal bleeding is a major complication of dual antiplatelet therapy (DAPT) in patients undergoing percutaneous coronary intervention (PCI). Malignancy may be detected due to gastrointestinal bleeding, necessitating critical decisions regarding treatment selection and influencing patient prognosis. This study investigated the frequency of gastrointestinal malignancies, associated factors, and prognoses in patients undergoing perioperative PCI through upper and lower gastrointestinal endoscopy.

**Methods:** This single-center, retrospective, observational study included 501 Japanese patients who underwent initial PCI between January 2019 and January 2023. Of these patients, 393 who underwent perioperative upper and lower gastrointestinal endoscopy were evaluated for the presence of gastrointestinal malignancy.

**Results:** Of the total patients, 36% presented with acute coronary syndrome (ACS). Gastrointestinal malignancies were identified in 30 patients (8%), including 18 cases of colorectal cancer and eight cases of gastric cancer. No difference in the frequency of malignancies was observed between patients with ACS and chronic coronary syndrome (CCS) (p = 0.7398). Malignancies were significantly more common in patients with positive faecal immunochemical testing (FIT) (p < 0.0001); however, FIT did not detect all malignancies. The 1500-day survival rate for patients with gastrointestinal malignancies was 64%, with no difference in overall survival between treatment modalities.

**Conclusions:** A considerable proportion of Japanese patients undergoing PCI had gastrointestinal malignancies, regardless of whether they had ACS or CCS, and their prognosis was poor. Upper and lower gastrointestinal endoscopic evaluations should be conducted, including for ACS cases, to prevent DAPT-induced gastrointestinal bleeding and improve prognosis.

## Introduction

The incidence of cardiovascular diseases is increasing worldwide owing to population growth and aging societies. Among these, ischemic heart disease (IHD) is the most common and a leading cause of death.^1,2^ Recent advances in treatment, particularly percutaneous coronary intervention (PCI), have reduced mortality rates; however, dual antiplatelet therapy (DAPT) is required as a standard drug therapy during the perioperative period of PCI. Since DAPT significantly reduces the risk of stent thrombosis but increases the risk of bleeding complications, multiple lines of evidence support a shorter duration of DAPT.^3–6^

Gastrointestinal bleeding is a major complication of DAPT that affects patient prognosis.^7^ Additionally, gastrointestinal bleeding is a trigger for malignancy detection during DAPT, necessitating critical decisions regarding treatment selection.

Faecal immunochemical testing (FIT) is a commonly used screening tool for gastrointestinal bleeding, particularly for colorectal cancer,^8,9^ and is often employed clinically to assess the risk of gastrointestinal bleeding before PCI. However, FIT has known limitations in detecting gastrointestinal malignancies.

Therefore, this study investigated the frequency of gastrointestinal malignancies, associated factors, and prognoses in patients undergoing perioperative PCI, utilizing upper and lower gastrointestinal endoscopy.

## Methods

### Study protocol and population

This was a single-center, retrospective, observational study. Some patient data from this study have recently been submitted (Higashino M, MD, PhD, unpublished data, 2024); however, this study aimed to demonstrate the utility of perioperative upper and lower gastrointestinal endoscopy in patients undergoing PCI, specifically to evaluate differences in the frequency of gastrointestinal malignancies in acute coronary syndrome (ACS) and chronic coronary syndrome (CCS) and to asses prognosis after malignancy detection.

A total of 501 hospitalized Japanese patients who underwent initial PCI for IHD, including ACS, were screened between January 2019 and January 2023. Of these, 108 patients who did not undergo either upper or lower gastrointestinal endoscopy during the perioperative period of PCI were excluded. The perioperative period was defined as early post-PCI during hospitalization for patients with ACS, and within 12 months before PCI for patients with CCS. Consequently, 393 patients were included in the final analysis (**Figure 1**). Patient information and clinical course data were collected from the hospital database. The study protocol was approved by the Institutional Review Board of National Hospital Organization Hakodate Medical Center (No.1105002), and patients were informed of their right to opt out of participation via the hospital’s homepage.

**Figure 1.**
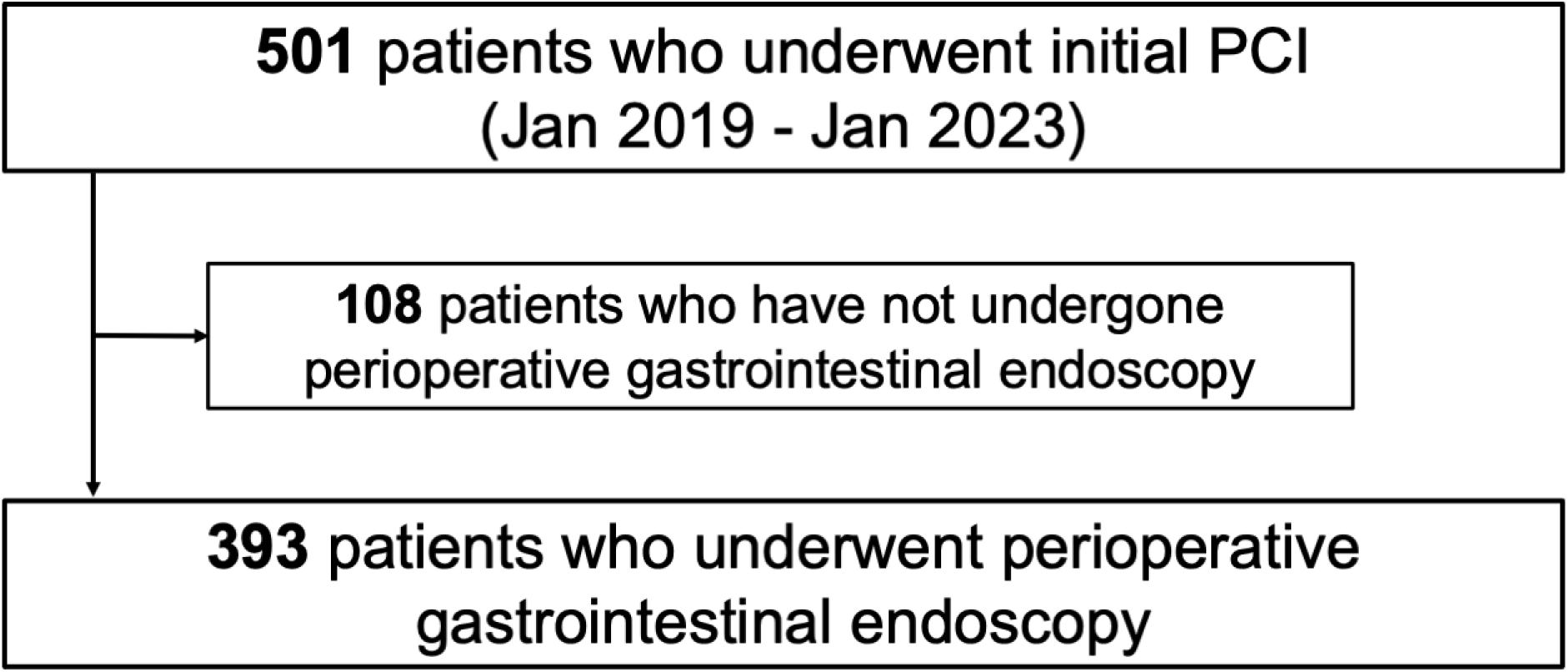
Patient selection. PCI, percutaneous coronary intervention.

### DAPT administration and gastrointestinal examination

All patients with ACS received a loading dose of DAPT prior to emergency revascularization, and the maintenance dose of DAPT was continued the following day. FIT and upper or lower gastrointestinal endoscopy were performed during hospitalization for the detection of haemorrhagic lesions or malignancies in the upper or lower gastrointestinal tract.

In patients with CCS, the dosage and timing of the antiplatelet agents, as well as the timing of FIT and endoscopy, were left to the discretion of the attending physicians. All patients were prescribed the appropriate doses of aspirin and prasugrel for DAPT.

### Statistical analysis

The Shapiro-Wilk test was used to assess the normality of continuous variables. Continuous variables are expressed as mean ± standard deviation or as median and interquartile range, and comparisons were made using either an unpaired Student’s t-test or a Wilcoxon rank-sum test, as appropriate. Categorical variables are expressed as numbers (percentages) and were compared between groups using the chi-square test.

To identify the independent determinants of malignancy detection, multivariate logistic regression analyses were performed using variables with a p-value < 0.05 in the univariable analyses. Kaplan-Meier analysis and the log-rank test were performed to assess overall survival. A p-value < 0.05 was considered significant for all tests. All statistical analyses were performed using JMP software (version 17.0; SAS Institute Inc., Cary, NC, USA).

## Results

### Patient characteristics

A total of 501 patients were screened, and 393 were enrolled in this study, as shown in **Figure 1**. The patient characteristics are summarized in **Table 1**. The age of the patients was 75 (66-82) years, and 277 patients (70 %) were male. Among the cohort, 36% had ACS, and 23% had heart failure. More than half of patients had a history of smoking. All patients underwent revascularization using drug-eluting stents. None of the patients had been treated or diagnosed with gastrointestinal malignancies prior to the targeted endoscopy.

**Table 1.** Patient characteristics.

| Variables | n=393 |
| --- | --- |
| Age, year | 75 (66 - 82) |
| Male, n (%) | 277 (70) |
| Body surface area, m<sup>2</sup> | 1.64 ± 0.19 |
| Acute coronary syndrome, n (%) | 142 (36) |
| Heart failure, n (%) | 89 (23) |
| Hypertension, n (%) | 299 (76) |
| Diabetes mellitus, n (%) | 159 (40) |
| Dyslipidemia, n (%) | 227 (58) |
| Atrial fibrillation / flutter, n (%) | 68 (17) |
| Smoking, n (%) | 219 (56) |
| Hemoglobin, g/dL | 12.7 ± 1.9 |
| Albumin, g/dL | 3.7 (3.4 - 4.0) |
| Sodium, mEq/L | 141 (139 - 143) |
| eGFR, mL/min/1.73m<sup>2</sup> | 58.9 (45.7 - 72.8) |
| BNP, pg/mL | 54.6 (21.7 - 154.3) |

Data are expressed as mean ± standard deviation, median (IQR), or n (%).
eGFR, estimated glomerular filtration rate; BNP, brain natriuretic peptide.

### Association between patients with PCI and malignancy

Gastrointestinal malignancies were found in 30 patients (8%), including 18 cases of colorectal cancer, eight cases of gastric cancer, two cases of esophageal cancer, one case of laryngeal cancer, and one overlapping case of gastric and colorectal cancer. Twelve patients were treated endoscopically, and 11 were treated surgically. There was no difference in the frequency of malignancy between patients with ACS and CCS (p = 0.7398) (**Figure 2**). FIT was performed in 177 cases without dietary or medicinal restrictions prior to endoscopy, and malignancies were significantly more frequent in cases with positive FIT results (p < 0.0001) (**Figure 3**). The sensitivity and specificity of FIT for detecting malignancy were 50% and 92%, respectively.

**Figure 2.**
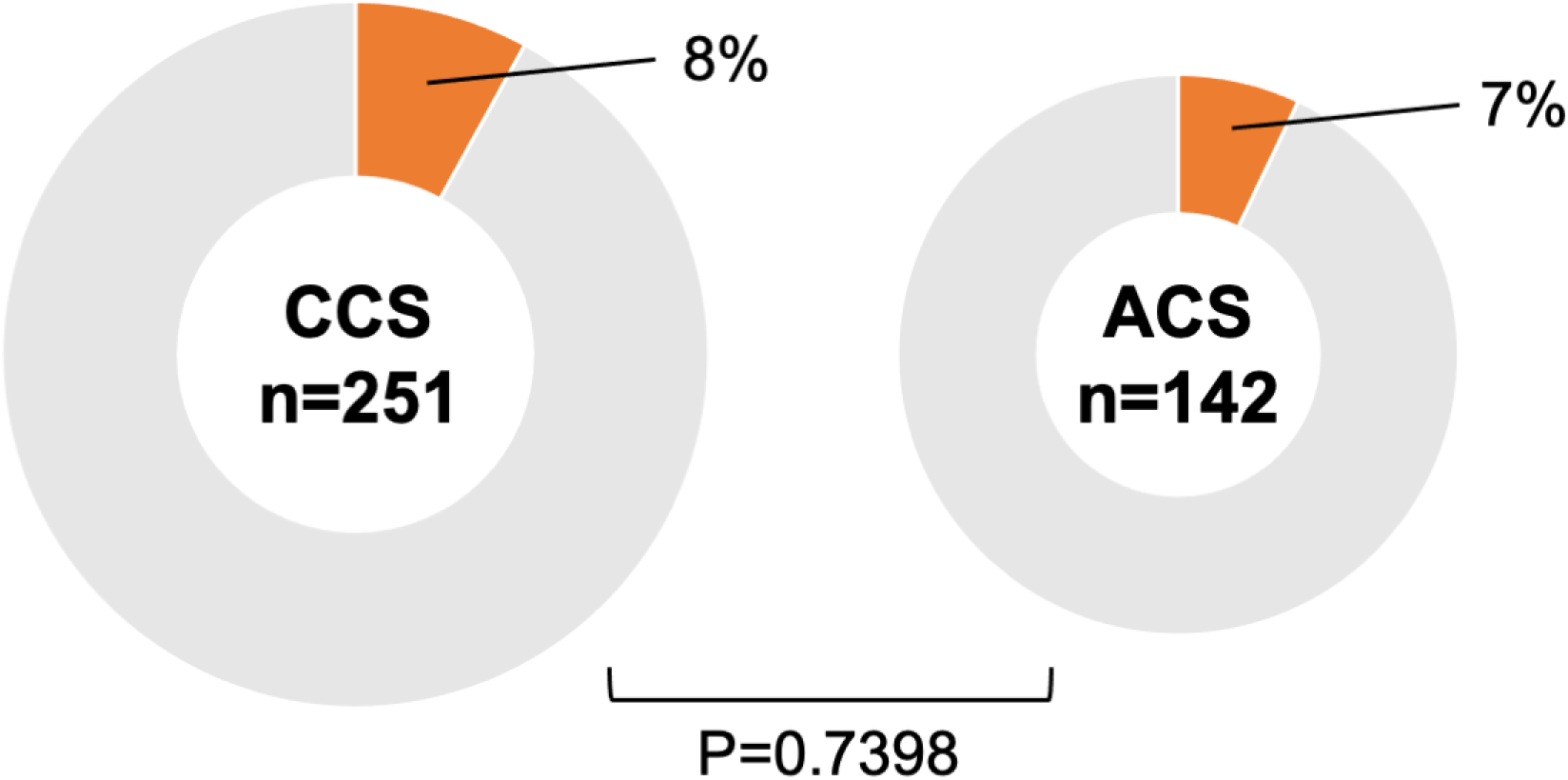
Association between ACS/CCS and malignancy. There was no difference in the frequency of malignancy between patients with ACS and CCS. ACS, acute coronary syndrome; CCS, chronic coronary syndrome.

**Figure 3.**
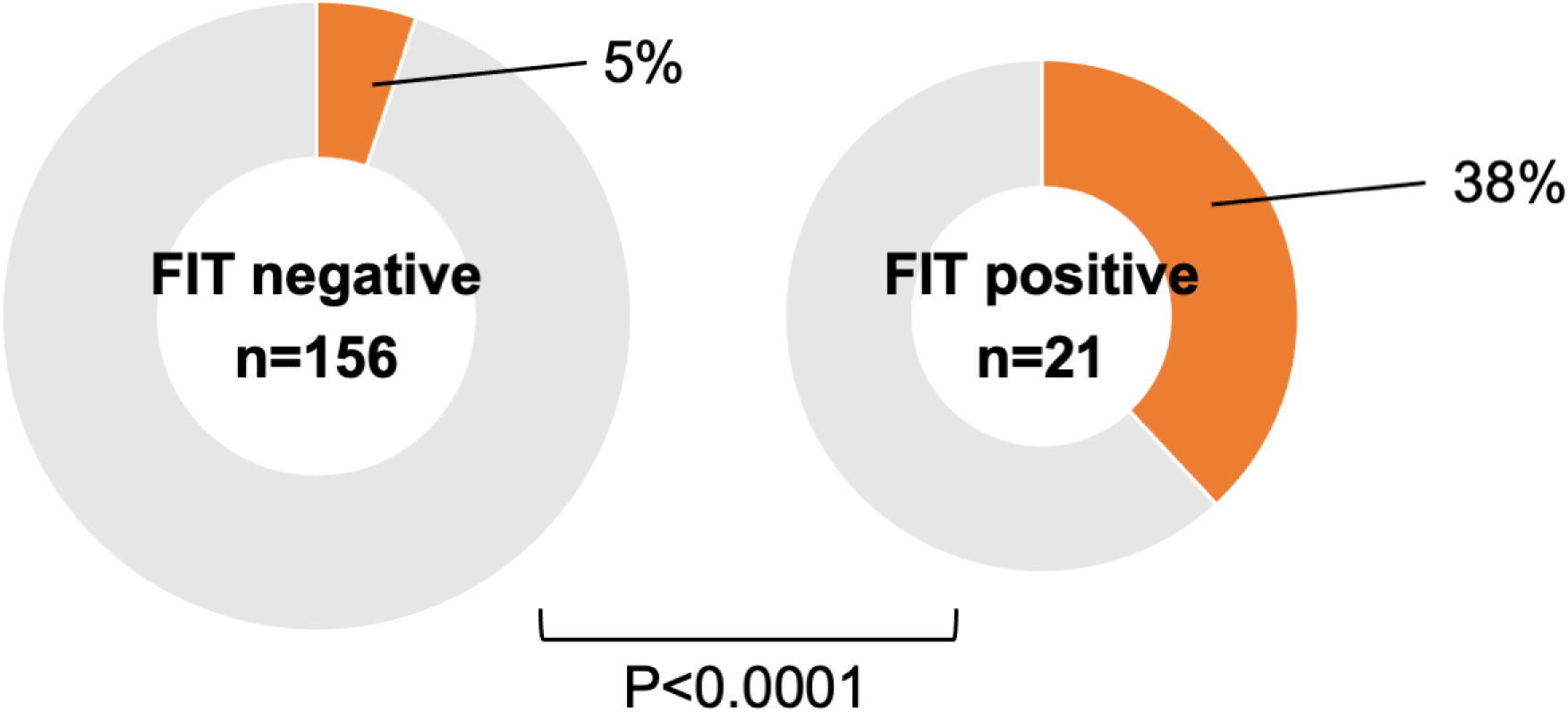
Association between FIT and malignancy. Gastrointestinal malignancies were significantly more frequent in cases with positive FIT. FIT, faecal immunochemical testing.

The association between malignancy and each parameter is shown in **Figure 4**. Age, haemoglobin level, and smoking history were not associated with malignancy; however, albumin level and FIT were significantly associated with malignancy.

**Figure 4.**
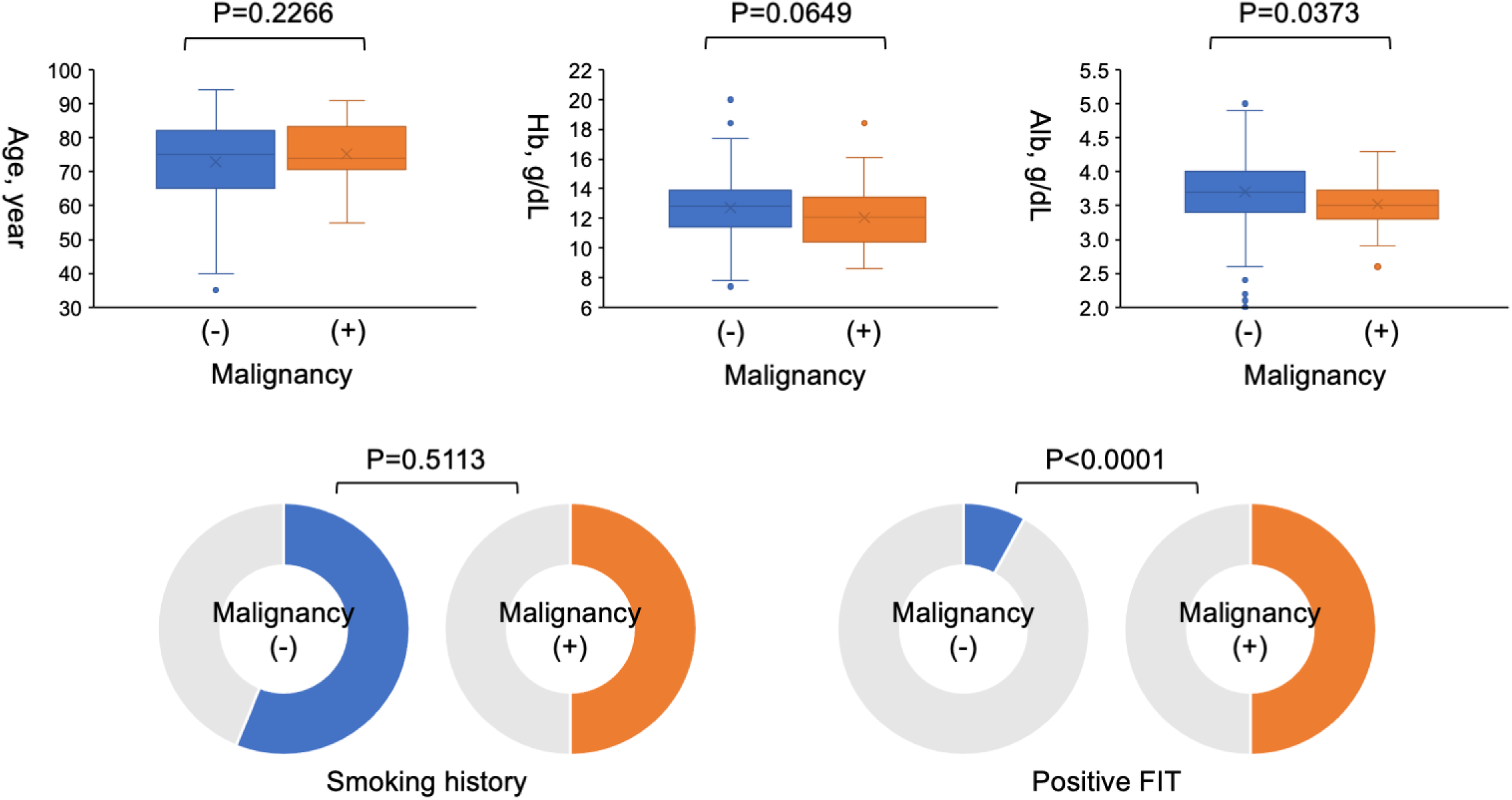
Association of each parameter with malignancy. Albumin level and FIT were significantly associated with malignancy. FIT, faecal immunochemical testing.

In the multivariate analysis for malignancy detection, only FIT results were significantly associated with malignancy (**Table 2**).

**Table 2.**
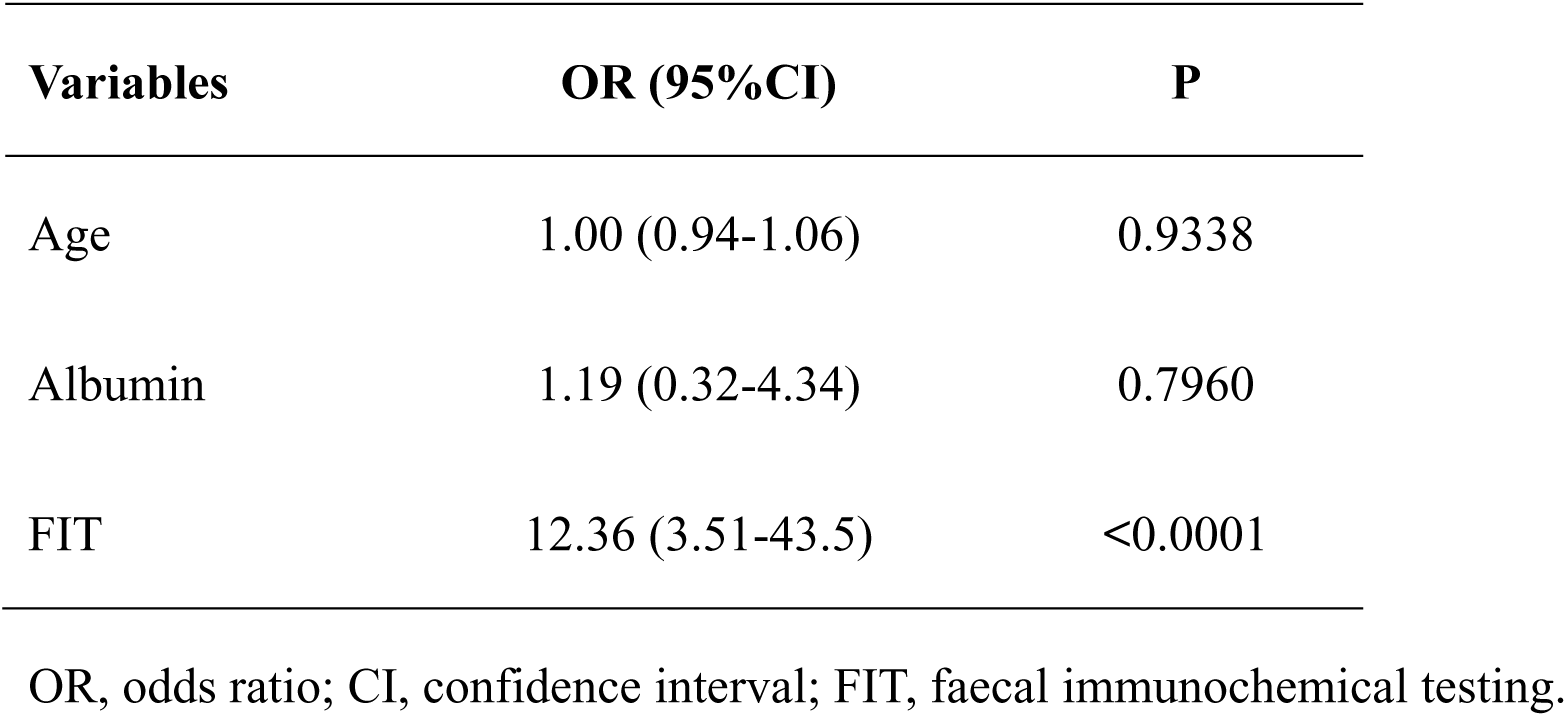
Multivariate analysis for malignancy detection.

| Variables | OR (95%CI) | P |
| --- | --- | --- |
| Age | 1.00 (0.94-1.06) | 0.9338 |
| Albumin | 1.19 (0.32-4.34) | 0.7960 |
| FIT | 12.36 (3.51-43.5) | <0.0001 |

OR, odds ratio; CI, confidence interval; FIT, faecal immunochemical testing.

### Prognosis of patients with PCI with gastrointestinal malignancies

During a median follow-up period of 1260 (796-1500) days, 10 patients with gastrointestinal malignancies died. The causes of death were myocardial infarction in one case, gastrointestinal malignancy in four cases, cancer treatment-related infection in two cases, anaemia in one case, and unknown causes in two cases.

Figure 5 shows the survival curves for patients with detected gastrointestinal malignancies. The survival rate at 1500 days was 64% for patients with gastrointestinal malignancies detected in the perioperative period of PCI. There was no difference in overall survival rate among the surgical, endoscopic, and other treatment groups (chemotherapy, radiation therapy, and best supportive care).

**Figure 5.**
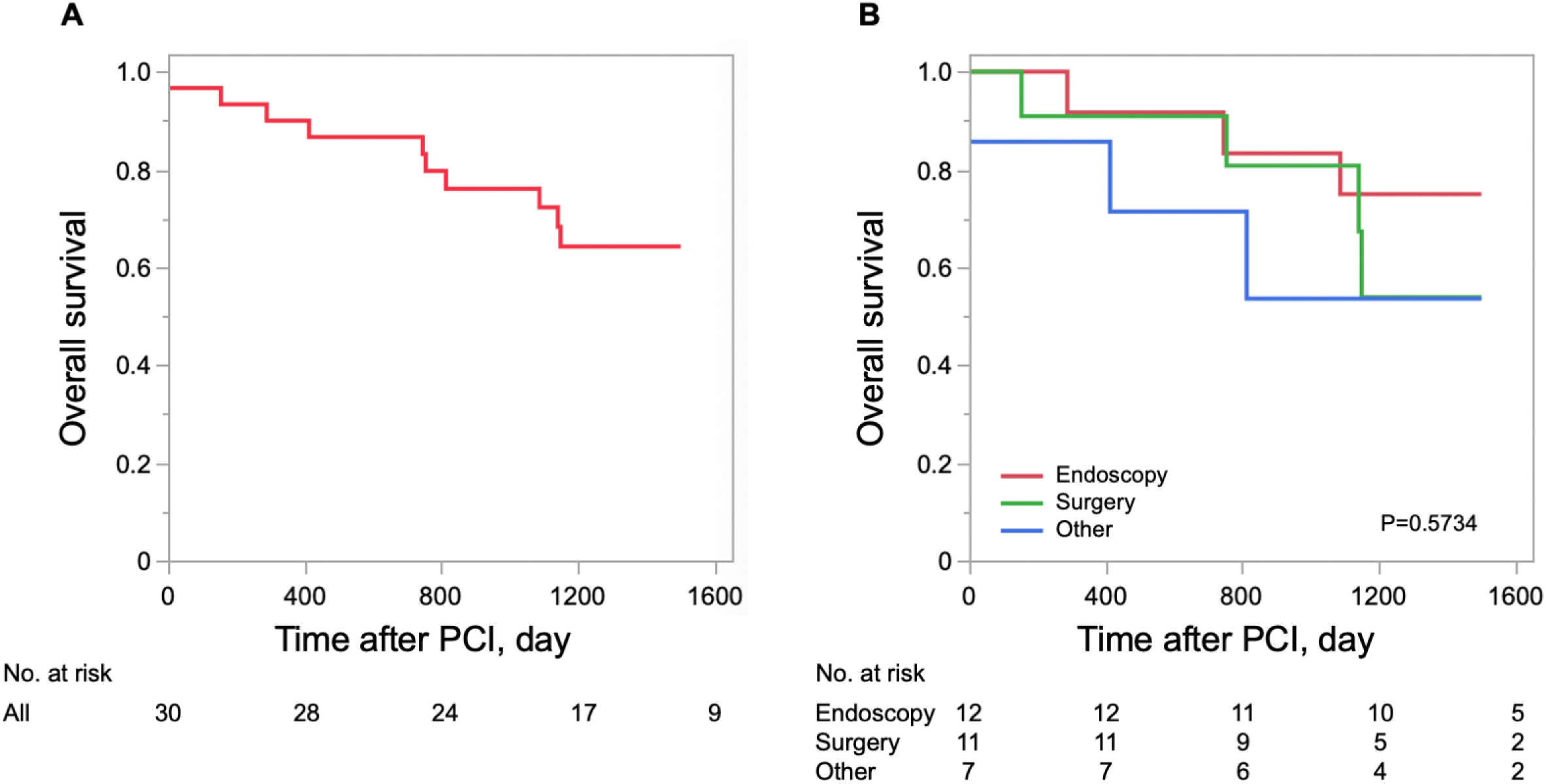
Kaplan-Meier curves for overall survival. (A) Survival curves for all patients with gastrointestinal malignancies. The survival rate at 1500 days was 64% in patients with gastrointestinal malignancies. (B) Survival curves for patients with malignancies by treatment. There was no difference in overall survival rate among the surgical, endoscopic, and other treatment groups (chemotherapy, radiation therapy, and best supportive care).

## Discussion

The main findings of this study are as follows: (1) gastrointestinal malignancies were present in 8% of PCI cases, and their prognosis was poor; (2) there was no difference in the frequency of gastrointestinal malignancies between patients with ACS and CCS; (3) positive FIT results were associated with the presence of gastrointestinal malignancy, but it did not detect all patients with malignancy. This was a clinically relevant study that performed both upper and lower gastrointestinal endoscopy in the perioperative period of PCI, whether for ACS or CCS, and examined the presence of gastrointestinal malignancies.

### Coronary artery disease and gastrointestinal cancer

Cardiovascular disease and cancer remain the two leading causes of death in developed countries, despite advances in their diagnosis, prevention, and treatment. Importantly, several classical risk factors and underlying pathophysiological mechanisms associated with cardiovascular disease are also linked to an increased risk of cancer.^10^ Chan et al. reported that 4.4% of patients with at least 50% stenosis in the major coronary arteries had colorectal cancer, significantly more than in the general population, and that metabolic syndrome and smoking were associated with advanced colorectal lesions and coronary artery disease.^11^ On the other hand, few reports have clarified the relationship between gastric cancer and IHD; however, it has been suggested that *Helicobacter pylori*, which induces a high rate of gastric cancer, may also induce an inflammatory response that may lead to coronary artery disease.^12,13^

From a surgical standpoint, Wilson et al. reported that 8.3% of patients who required non-cardiac surgery within 60 days of coronary stenting underwent gastrointestinal or abdominal surgery.^14^ Tokushige et al. reported that patients undergoing PCI frequently underwent abdominal surgery or gastrointestinal endoscopic procedures within a few years.^15^

Thus, this suggest that there is a significant association between gastrointestinal malignancies and coronary artery disease.

### Non-cardiac surgery and coronary revascularization

The risk of cardiovascular events in the perioperative period of non-cardiac surgery increases in the presence of IHD.^16,17^ However, prophylactic revascularization before major vascular surgery for all patients with stable coronary disease does not improve the perioperative period or the long-term clinical course.^18,19^ In addition, preoperative revascularization in patients with stable IHD before noncardiac surgery is not beneficial, as it does not reduce perioperative mortality or the incidence of acute myocardial infarction.^20^ Regardless of whether IHD or malignancy is detected earlier, it is reasonable to assume that if invasive treatment for malignancy is required, it should be prioritized before revascularization for stable IHD.

Although ACS is preceded by revascularization, the risk of cardiovascular events in the perioperative period of non-cardiac surgery is particularly high within the first month after onset and decreases over time.^21–25^ Surgical delays are desirable when malignancy is detected shortly after myocardial infarction; however, appropriate surgery may still be feasible with adequate discussion between the cardiologist and oncologist. Although cardiac death accounts for the majority of fatalities in the first month after the onset of ACS, non-cardiac disease accounts for two-thirds of deaths, with malignancy accounting for a particularly large proportion.^26^ The detection of malignancy is also important in patients with ACS since long-term prognosis is often determined by non-cardiac disease.

### Antiplatelet therapy and bleeding complications

Since the efficacy of DAPT in preventing stent thrombosis has been established, it is now the standard of care after PCI.^27^ However, antiplatelet therapy carries a risk of increased bleeding complications, and DAPT is associated with a 7.4-fold increase in the risk of upper gastrointestinal bleeding.^28^

Active malignancies are one of the high-risk factors for bleeding.^29–31^ Gastric and colorectal cancers are the major causes of cancer-related death by organ,^32^ and these gastrointestinal malignancies are often clinically detected following gastrointestinal bleeding during DAPT. The risk of stent thrombosis may increase during endoscopic or surgical treatment of gastrointestinal malignancies, which may require DAPT withdrawal. In this context, it is reasonable to evaluate for malignancy before PCI in CCS. Even if DAPT has already been introduced after ACS, endoscopy without biopsy does not necessitate DAPT withdrawal.^33^ Considering the aforementioned risk of bleeding complications, performing endoscopy to determine the treatment strategy is advisable, even in patients with ACS.

### FIT and malignancy

FIT is a well-established strategy for colorectal cancer screening in the general population.^8,9^ The detection rate of colorectal cancer by FIT is comparable to that of endoscopy,^9^ and should be favoured as an initial test due to its simplicity and noninvasive nature. On the other hand, FIT may yield positive results for upper gastrointestinal haemorrhage only in cases of massive haemorrhage. However, the positive rate of FIT is low because haemoglobin is often degraded by proteases in gastric and pancreatic juices or denatured by other digestive juices, reducing its antigenic properties. Although the usefulness of FIT for detecting upper gastrointestinal bleeding remains controversial, the diagnostic significance of performing upper and lower gastrointestinal endoscopy during the perioperative period of PCI with DAPT is evident, considering that cancers at risk for bleeding can be detected even in FIT-negative cases, and that it was possible to detect both gastric and colorectal cancers in this study.

### Clinical Significance of this Study

This study is the first to investigate the prevalence and factors of malignancy through upper and lower gastrointestinal endoscopy in PCI’s perioperative period. In patients with ACS, endoscopy was performed promptly after acute revascularization one hemodynamic stability was achieved. Early malignancy detection allows timely discussions between cardiologists and oncologists for treatment planning. Patients with CCS, treated on standby basis, have the advantage of undergoing endoscopy before coronary intervention, facilitating early management of malignancy or haemorrhagic lesions. No complications were associated with endoscopy.

Although FIT provides an opportunity to detect malignancies, it does not identify all malignancies or gastrointestinal bleeding. Therefore, gastrointestinal endoscopy should be performed whenever possible, including in ACS cases. Notably, despite detection in the perioperative period, malignancy prognosis remains poor, irrespective of treatment.

### Limitations

First, this was a single-center, retrospective, observational study with a relatively small sample size. Therefore, the number of patients with malignancy was limited, and many variables could not be entered into the final multivariable models. Second, the timing of DAPT and gastrointestinal evaluation was left to the discretion of the attending physician in line with clinical practice, and the association between antiplatelet drug administration and malignancy detection was not clear. In addition, FIT was not performed in many cases, particularly in CCS cases. Third, although gastric cancer was clearly associated with *Helicobacter pylori* carriage, we were unable to assess the presence of *Helicobacter pylori* carriage in all patients in our study. Furthermore, tumour marker measurements that suggested the presence of gastrointestinal malignancies were not performed.

## Conclusions

A high proportion of Japanese patients undergoing PCI had gastrointestinal malignancies, regardless of whether they had ACS or CCS, and their prognosis was poor. Gastrointestinal endoscopic evaluation is essential for accurate malignancy diagnosis, prevention of DAPT-induced gastrointestinal bleeding, and improving prognosis, including in ACS cases.

## Data Availability

We have full access to all study data referred to in the manuscript.

## Non-standard Abbreviations and Acronyms

ACS: acute coronary syndrome
CCS: chronic coronary syndrome
DAPT: dual antiplatelet therapy
FIT: faecal immunochemical testing
IHD: ischemic heart disease
PCI: percutaneous coronary intervention

## Acknowledgements

We thank Editage (www.editage.com) for English language editing. This study received no specific grant from any funding agency.

---

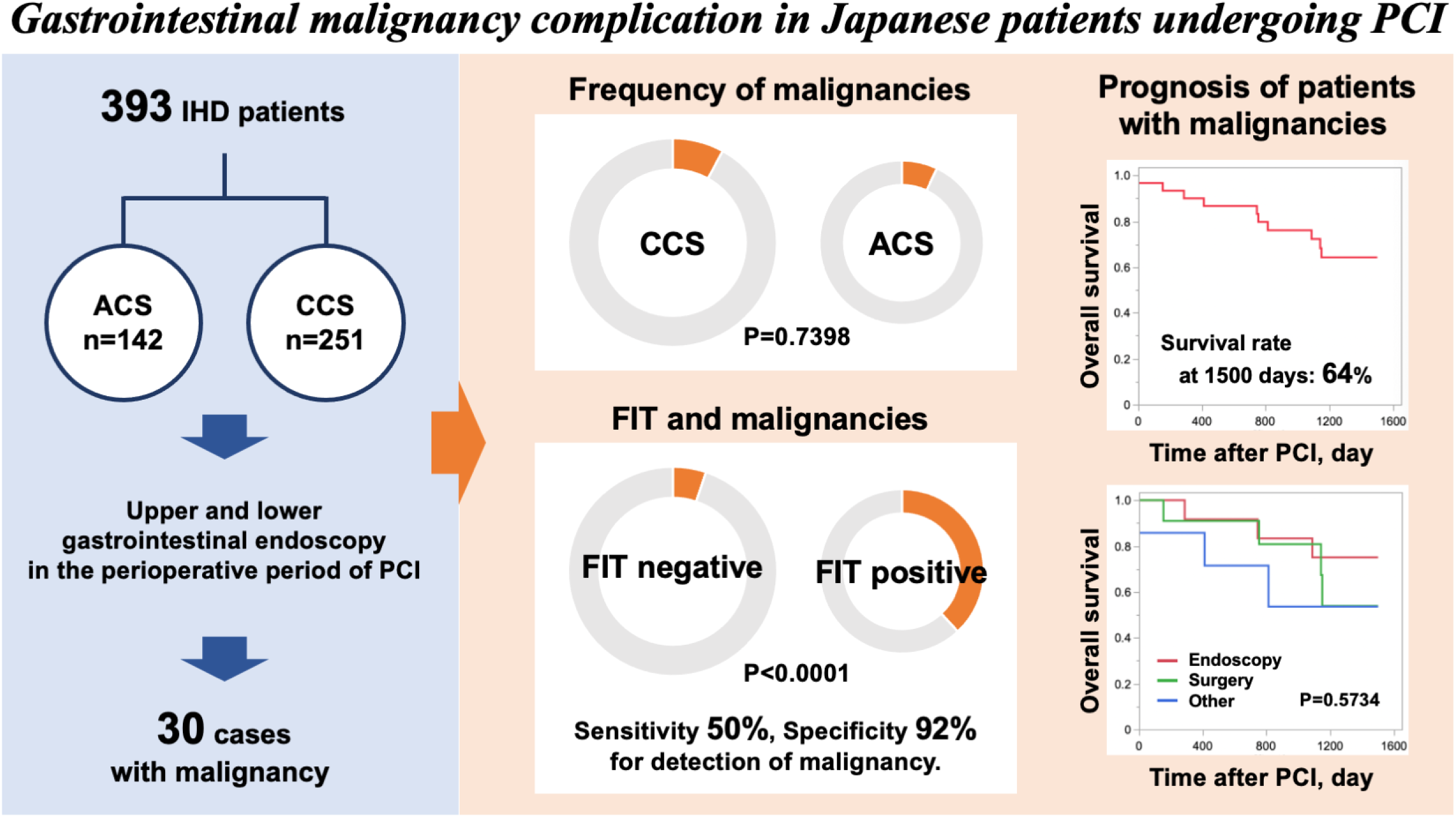

## References

1. Roth GA, Mensah GA, Fuster V. The Global Burden of Cardiovascular Diseases and Risks: A Compass for Global Action. J Am Coll Cardiol. 2020;76:2980–2981. doi: 10.1016/j.jacc.2020.11.021

2. Vaduganathan M, Mensah GA, Turco JV, Fuster V, Roth GA. The Global Burden of Cardiovascular Diseases and Risk: A Compass for Future Health. J Am Coll Cardiol. 2022;80:2361–2371. doi: 10.1016/j.jacc.2022.11.005

3. Palmerini T, Benedetto U, Bacchi-Reggiani L, Della Riva D, Biondi-Zoccai G, Feres F, Abizaid A, Hong MK, Kim BK, Jang Y, et al. Mortality in patients treated with extended duration dual antiplatelet therapy after drug-eluting stent implantation: a pairwise and Bayesian network meta-analysis of randomised trials. Lancet. 2015;385:2371–2382. doi: 10.1016/s0140-6736(15)60263-x

4. Valgimigli M, Frigoli E, Heg D, Tijssen J, Jüni P, Vranckx P, Ozaki Y, Morice MC, Chevalier B, Onuma Y, et al. Dual Antiplatelet Therapy after PCI in Patients at High Bleeding Risk. N Engl J Med. 2021;385:1643–1655. doi: 10.1056/NEJMoa2108749

5. Watanabe H, Domei T, Morimoto T, Natsuaki M, Shiomi H, Toyota T, Ohya M, Suwa S, Takagi K, Nanasato M, et al. Effect of 1-Month Dual Antiplatelet Therapy Followed by Clopidogrel vs 12-Month Dual Antiplatelet Therapy on Cardiovascular and Bleeding Events in Patients Receiving PCI: The STOPDAPT-2 Randomized Clinical Trial. Jama. 2019;321:2414–2427. doi: 10.1001/jama.2019.8145

6. Toyota T, Shiomi H, Morimoto T, Natsuaki M, Kimura T. Short versus prolonged dual antiplatelet therapy (DAPT) duration after coronary stent implantation: A comparison between the DAPT study and 9 other trials evaluating DAPT duration. PLoS One. 2017;12:e0174502. doi: 10.1371/journal.pone.0174502

7. Nikolsky E, Stone GW, Kirtane AJ, Dangas GD, Lansky AJ, McLaurin B, Lincoff AM, Feit F, Moses JW, Fahy M, et al. Gastrointestinal bleeding in patients with acute coronary syndromes: incidence, predictors, and clinical implications: analysis from the ACUITY (Acute Catheterization and Urgent Intervention Triage Strategy) trial. J Am Coll Cardiol. 2009;54:1293–1302. doi: 10.1016/j.jacc.2009.07.019

8. Helsingen LM, Vandvik PO, Jodal HC, Agoritsas T, Lytvyn L, Anderson JC, Auer R, Murphy SB, Almadi MA, Corley DA, et al. Colorectal cancer screening with faecal immunochemical testing, sigmoidoscopy or colonoscopy: a clinical practice guideline. Bmj. 2019;367:l5515. doi: 10.1136/bmj.l5515

9. Quintero E, Castells A, Bujanda L, Cubiella J, Salas D, Lanas Á, Andreu M, Carballo F, Morillas JD, Hernández C, et al. Colonoscopy versus fecal immunochemical testing in colorectal-cancer screening. N Engl J Med. 2012;366:697–706. doi: 10.1056/NEJMoa1108895

10. Handy CE, Quispe R, Pinto X, Blaha MJ, Blumenthal RS, Michos ED, Lima JAC, Guallar E, Ryu S, Cho J, et al. Synergistic Opportunities in the Interplay Between Cancer Screening and Cardiovascular Disease Risk Assessment: Together We Are Stronger. Circulation. 2018;138:727–734. doi: 10.1161/circulationaha.118.035516

11. Chan AO, Jim MH, Lam KF, Morris JS, Siu DC, Tong T, Ng FH, Wong SY, Hui WM, Chan CK, et al. Prevalence of colorectal neoplasm among patients with newly diagnosed coronary artery disease. Jama. 2007;298:1412–1419. doi: 10.1001/jama.298.12.1412

12. Sharma V, Aggarwal A. Helicobacter pylori: Does it add to risk of coronary artery disease. World J Cardiol. 2015;7:19–25. doi: 10.4330/wjc.v7.i1.19

13. Li B, Zhang Y, Zheng Y, Cai H. The causal effect of Helicobacter pylori infection on coronary heart disease is mediated by the body mass index: a Mendelian randomization study. Scientific Reports. 2024;14:1688. doi: 10.1038/s41598-024-51701-8

14. Wilson SH, Fasseas P, Orford JL, Lennon RJ, Horlocker T, Charnoff NE, Melby S, Berger PB. Clinical outcome of patients undergoing non-cardiac surgery in the two months following coronary stenting. J Am Coll Cardiol. 2003;42:234–240. doi: 10.1016/s0735-1097(03)00622-3

15. Tokushige A, Shiomi H, Morimoto T, Furukawa Y, Nakagawa Y, Kadota K, Iwabuchi M, Shizuta S, Tada T, Tazaki J, et al. Incidence and outcome of surgical procedures after coronary bare-metal and drug-eluting stent implantation: a report from the CREDO-Kyoto PCI/CABG registry cohort-2. Circ Cardiovasc Interv. 2012;5:237–246. doi: 10.1161/circinterventions.111.963728

16. Koshy AN, Ha FJ, Gow PJ, Han HC, Amirul-Islam FM, Lim HS, Teh AW, Farouque O. Computed tomographic coronary angiography in risk stratification prior to non-cardiac surgery: a systematic review and meta-analysis. Heart. 2019;105:1335–1342. doi: 10.1136/heartjnl-2018-314649

17. Eagle KA, Coley CM, Newell JB, Brewster DC, Darling RC, Strauss HW, Guiney TE, Boucher CA. Combining clinical and thallium data optimizes preoperative assessment of cardiac risk before major vascular surgery. Ann Intern Med. 1989;110:859–866. doi: 10.7326/0003-4819-110-11-859

18. McFalls EO, Ward HB, Moritz TE, Goldman S, Krupski WC, Littooy F, Pierpont G, Santilli S, Rapp J, Hattler B, et al. Coronary-artery revascularization before elective major vascular surgery. N Engl J Med. 2004;351:2795–2804. doi: 10.1056/NEJMoa041905

19. Garcia S, Moritz TE, Ward HB, Pierpont G, Goldman S, Larsen GC, Littooy F, Krupski W, Thottapurathu L, Reda DJ, et al. Usefulness of revascularization of patients with multivessel coronary artery disease before elective vascular surgery for abdominal aortic and peripheral occlusive disease. Am J Cardiol. 2008;102:809–813. doi: 10.1016/j.amjcard.2008.05.022

20. Wong EY, Lawrence HP, Wong DT. The effects of prophylactic coronary revascularization or medical management on patient outcomes after noncardiac surgery--a meta-analysis. Can J Anaesth. 2007;54:705–717. doi: 10.1007/bf03026867

21. Livhits M, Ko CY, Leonardi MJ, Zingmond DS, Gibbons MM, de Virgilio C. Risk of surgery following recent myocardial infarction. Ann Surg. 2011;253:857–864. doi: 10.1097/SLA.0b013e3182125196

22. Hawn MT, Graham LA, Richman JS, Itani KM, Henderson WG, Maddox TM. Risk of major adverse cardiac events following noncardiac surgery in patients with coronary stents. Jama. 2013;310:1462–1472. doi: 10.1001/jama.2013.278787

23. Holcomb CN, Hollis RH, Graham LA, Richman JS, Valle JA, Itani KM, Maddox TM, Hawn MT. Association of Coronary Stent Indication With Postoperative Outcomes Following Noncardiac Surgery. JAMA Surg. 2016;151:462–469. doi: 10.1001/jamasurg.2015.4545

24. Tokushige A, Shiomi H, Morimoto T, Ono K, Furukawa Y, Nakagawa Y, Kadota K, Iwabuchi M, Shizuta S, Tada T, et al. Influence of initial acute myocardial infarction presentation on the outcome of surgical procedures after coronary stent implantation: a report from the CREDO-Kyoto PCI/CABG Registry Cohort-2. Cardiovasc Interv Ther. 2013;28:45–55. doi: 10.1007/s12928-012-0136-x

25. Livhits M, Gibbons MM, de Virgilio C, O’Connell JB, Leonardi MJ, Ko CY, Zingmond DS. Coronary revascularization after myocardial infarction can reduce risks of noncardiac surgery. J Am Coll Surg. 2011;212:1018–1026. doi: 10.1016/j.jamcollsurg.2011.02.018

26. Pedersen F, Butrymovich V, Kelbæk H, Wachtell K, Helqvist S, Kastrup J, Holmvang L, Clemmensen P, Engstrøm T, Grande P, et al. Short- and long-term cause of death in patients treated with primary PCI for STEMI. J Am Coll Cardiol. 2014;64:2101–2108. doi: 10.1016/j.jacc.2014.08.037

27. Leon MB, Baim DS, Popma JJ, Gordon PC, Cutlip DE, Ho KK, Giambartolomei A, Diver DJ, Lasorda DM, Williams DO, et al. A clinical trial comparing three antithrombotic-drug regimens after coronary-artery stenting. Stent Anticoagulation Restenosis Study Investigators. N Engl J Med. 1998;339:1665–1671. doi: 10.1056/nejm199812033392303

28. Hallas J, Dall M, Andries A, Andersen BS, Aalykke C, Hansen JM, Andersen M, Lassen AT. Use of single and combined antithrombotic therapy and risk of serious upper gastrointestinal bleeding: population based case-control study. Bmj. 2006;333:726. doi: 10.1136/bmj.38947.697558.AE

29. Urban P, Mehran R, Colleran R, Angiolillo DJ, Byrne RA, Capodanno D, Cuisset T, Cutlip D, Eerdmans P, Eikelboom J, et al. Defining High Bleeding Risk in Patients Undergoing Percutaneous Coronary Intervention. Circulation. 2019;140:240–261. doi: 10.1161/circulationaha.119.040167

30. Natsuaki M, Morimoto T, Yamaji K, Watanabe H, Yoshikawa Y, Shiomi H, Nakagawa Y, Furukawa Y, Kadota K, Ando K, et al. Prediction of Thrombotic and Bleeding Events After Percutaneous Coronary Intervention: CREDO-Kyoto Thrombotic and Bleeding Risk Scores. J Am Heart Assoc. 2018;7. doi: 10.1161/jaha.118.008708

31. Ito S, Watanabe H, Morimoto T, Yoshikawa Y, Shiomi H, Shizuta S, Ono K, Yamaji K, Soga Y, Hyodo M, et al. Impact of Baseline Thrombocytopenia on Bleeding and Mortality After Percutaneous Coronary Intervention. Am J Cardiol. 2018;121:1304–1314. doi: 10.1016/j.amjcard.2018.02.010

32. Mabe K, Inoue K, Kamada T, Kato K, Kato M, Haruma K. Endoscopic screening for gastric cancer in Japan: Current status and future perspectives. Dig Endosc. 2022;34:412–419. doi: 10.1111/den.14063

33. Rossini R, Musumeci G, Visconti LO, Bramucci E, Castiglioni B, De Servi S, Lettieri C, Lettino M, Piccaluga E, Savonitto S, et al. Perioperative management of antiplatelet therapy in patients with coronary stents undergoing cardiac and non-cardiac surgery: a consensus document from Italian cardiological, surgical and anaesthesiological societies. EuroIntervention. 2014;10:38–46. doi: 10.4244/eijv10i1a8

